# A Community-Based Strategy to Improve Identification and Linkage of Pregnant Women to Antenatal and HIV Care Services in Gombe State, Nigeria

**DOI:** 10.64898/2026.02.10.26345966

**Authors:** Abdulkarim Suraj, Bappah Lawan, Abubakar Abdulrazaq Ahmad, Danimoh Mustapha Abdulsalam

## Abstract

**Background:** Nigeria accounts for a significant share of global maternal mortality, and HIV remains a public health threat. Gombe State in northeastern Nigeria contends with profound barriers to healthcare access. This study evaluated the effectiveness of a community-based intervention using trained Community Health Workers (CHWs) to improve early identification of pregnancy and linkage to Antenatal Care (ANC) and HIV services.

**Methods:** A quasi-experimental design was employed across six local government areas (LGAs) from January 2020 to June 2021. Three LGAs were randomly assigned to the intervention, where CHWs conducted home visits for pregnancy identification, health education, and referral facilitation. Three control LGAs received standard facility-based care. Data were collected via household surveys and facility records at baseline and endline. Analysis included Difference-in-Differences (DiD) estimation to determine the net intervention effect.

**Results:** The intervention group showed significant improvements compared to the control. Early pregnancy identification (<20 weeks) increased from 45% to 78% (DiD: +29 pp, p<0.001). Attendance of at least one ANC visit rose from 58% to 85% (DiD: +22 pp, p<0.001), reducing the coverage gap by 89%. Subgroup analysis revealed the largest gains among adolescents (DiD: +31 pp) and rural residents (DiD: +27 pp). HIV testing uptake increased from 52% to 90% (DiD: +34 pp, p<0.001). Linkage to care for HIV-positive women improved from 65% to 92% (p=0.002).

**Conclusion:** A CHW-led, community-based strategy is highly effective in improving early engagement with ANC and HIV services in resource-limited settings. The intervention demonstrated a strong equity-promoting effect. Integration and scale-up of this model within primary healthcare systems is recommended.

## 1. Introduction

Nigeria remains a critical focal point in the global effort to improve maternal health and combat the HIV epidemic. The country accounts for approximately 20% of global maternal deaths, with an estimated Maternal Mortality Ratio (MMR) of 512 per 100,000 live births a figure that has remained stubbornly high despite decades of targeted interventions (WHO, 2019). This crisis is compounded by the persistent threat of HIV, with a national prevalence of 1.4% among pregnant women and significant regional disparities that underscore the uneven distribution of healthcare access and prevention services (NACA, 2020). The co-existence of high maternal mortality and HIV burden creates a synergistic public health challenge, particularly in regions where health systems are fragile and socio-cultural barriers are pervasive.

Gombe State, situated in Nigeria’s North-East geopolitical zone, epitomizes this confluence of challenges. Characterized by a predominantly rural population, high poverty levels, and low female literacy, the state reports some of the nation’s most alarming health indicators. The state’s MMR is estimated at 1,549 per 100,000 live births, a rate that not only dwarfs the national average but also signals profound systemic failures in the provision and utilization of essential maternal healthcare services (Gombe SMOH, 2018). Concurrently, while national HIV prevalence has declined, Gombe and other northern states continue to experience localized epidemics and face significant hurdles in achieving the 95-95-95 targets for Prevention of Mother-to-Child Transmission (PMTCT) (UNAIDS, 2022). Nigeria still accounts for a large share of global new pediatric HIV infections, a reality driven in part by gaps in service uptake in regions like the North-East.

Timely initiation and consistent utilization of quality Antenatal Care (ANC) is universally recognized as a cornerstone for reducing maternal and perinatal mortality. It is the critical entry point for a bundle of life-saving interventions, including the prevention of vertical HIV transmission. However, the coverage and quality of ANC in Northern Nigeria remain severely inadequate. According to the 2018 Nigeria Demographic and Health Survey (NDHS), only 57% of pregnant women in the North-East received ANC from a skilled provider at least once, and a mere 15% completed the recommended minimum of four visits far below national averages of 67% and 44%, respectively. Furthermore, late initiation of care, often well into the second or third trimester, is commonplace. This delay deprives women of vital early interventions, including tetanus immunization, malaria prophylaxis, iron supplementation, and, crucially, early HIV testing and counseling (Adeyinka *et al*., 2020; Banke-Thomas *et al*., 2020).

A complex, multi-layered web of barriers underlies these poor outcomes. These barriers operate at individual, community, and health system levels:

i. **Geographical and Financial Barriers:** Distance to functional health facilities, poor transportation infrastructure, and direct and indirect costs (transport, service fees, informal payments) are primary deterrents, especially for rural women (Okonofua et al., 2017).
ii. **Socio-cultural and Gender Norms:** Deeply entrenched patriarchal structures often limit women’s autonomy and decision-making power regarding their own health. The need for spousal or familial permission to seek care, coupled with a preference for home births attended by Traditional Birth Attendants (TBAs), significantly restricts facility-based service utilization (Izugbara et al., 2018; Iliyasu et al., 2019).
iii. **Health System Weaknesses:** Chronic issues such as shortages of skilled personnel, stock-outs of essential drugs and test kits, long waiting times, and perceived or actual poor quality of care and negative staff attitudes erode community trust and discourage service uptake (Oleribe et al., 2016).
iv. **HIV-Related Stigma:** Fear of social ostracization, discrimination, blame, and even violence upon testing positive is a pervasive and powerful barrier. This stigma deters women from accepting HIV testing during ANC and complicates disclosure and linkage to care for those who test positive, with confidentiality concerns being particularly acute in close-knit communities (Turan *et al*., 2018).

In response to such multifaceted barriers, community-based strategies have emerged as a promising and evidence-based approach to bridge the gap between communities and formal health systems. Systematic reviews affirm that interventions deploying Community Health Workers (CHWs) for pregnancy identification, health education, and referral support can significantly increase the proportion of women initiating ANC early and attending the recommended number of visits (Lassi *et al*., 2015; Amoakoh-Coleman *et al*., 2015). CHWs, typically selected from and trusted by their own communities, can effectively overcome geographical and social distance. Their proximity allows for regular home visits, enabling early pregnancy detection and sustained engagement through pregnancy and the postpartum period (Perry & Zulliger, 2012). Moreover, community-based approaches are vital for improving PMTCT outcomes. CHWs can provide pre-test counseling, facilitate testing (including home-based testing where feasible), and offer post-test support and follow-up, which significantly increases HIV testing rates and timely ART initiation among pregnant women (Siegfried et al., 2014).

While the efficacy of community-based models is well-documented globally, their success in a specific context like Gombe State requires careful adaptation to local realities. This includes navigating patriarchal social structures, potential security concerns, and leveraging existing community networks such as Ward Development Committees and traditional leadership. Studies from similar northern Nigerian contexts highlight the critical importance of engaging religious and traditional leaders as champions and tailoring health messages to align with local cultural and religious norms for program acceptance and sustainability (Adebayo *et al*., 2018).

Despite this compelling evidence base, there remains a need for robust, locally-generated evidence on the implementation and impact of such integrated (ANC/HIV) community-based models in Gombe State. This study was therefore designed to evaluate the effectiveness of a tailored, CHW-led community-based intervention on enhancing the early identification of pregnant women and promoting their linkage to and utilization of ANC and HIV care services within the specific socio-cultural and health system context of Gombe State, Nigeria.

## Methods

### Study Setting

The study was conducted in Gombe State, Northeast Nigeria. The state has an estimated population of 4.1 million, characterized by ethnic diversity, a predominantly Muslim population in northern regions, high poverty levels, and low female literacy. Administrative divisions include eleven Local Government Areas (LGAs), with significant variation in terrain and health infrastructure between northern and southern zones.

### Study Design

A quasi-experimental study with a control group was implemented from January 2020 to June 2021. Six predominantly rural LGAs facing similar socio-economic and infrastructural challenges were selected. Using a random assignment procedure, three LGAs (Kwami, Kaltungo, Akko) were allocated to the intervention arm, and three (Dukku, Billiri, Yamaltu Deba) served as controls.

### Study Population

The target population was pregnant women aged 15-49 years residing in the selected LGAs.

### Intervention Description

The intervention comprised several key components:

1. **Training:** 120 CHWs were trained on pregnancy identification, the importance of early ANC and HIV testing, counseling techniques, and referral procedures.
2. **Community Activity:** CHWs conducted monthly house-to-house visits to identify pregnant women, provide tailored health education, and encourage facility registration. They offered appointment reminders and, when necessary, escorted women to health facilities.
3. **Health System Linkage:** Basic strengthening of receiving health facilities was conducted to ensure readiness and friendly service delivery for referred clients. Control LGAs continued with routine, passive health facility-based services without active CHW outreach.

### Data Collection and Sources

Data were collected at baseline (pre-intervention) and endline (post-intervention).

1. **Household Surveys:** A multi-stage cluster sampling technique was used to administer structured questionnaires to 400 women per study arm at each time point. Surveys captured socio-demographics, ANC attendance, timing of first visit, HIV testing history, and perceived barriers.
2. **Routine Health Facility Data:** ANC registers, PMTCT logs, and ART registers were reviewed to track service utilization and the HIV care cascade.
3. **Program Monitoring Data:** CHW activity registers and referral tracking forms were used to monitor intervention fidelity.

### Data Analysis

Data were analyzed using SPSS software. Descriptive statistics summarized participant characteristics. Chi-square tests assessed baseline comparability between groups. The primary analysis utilized Difference-in-Differences (DiD) estimation to isolate the net effect of the intervention on primary outcomes: 1) Early pregnancy identification (<20 weeks), 2) Attendance of at least one ANC visit, 3) HIV testing uptake, and 4) Linkage to care for HIV-positive women. Subgroup analysis was performed for ANC attendance.

### Ethical Considerations

Ethical approval was obtained from the Gombe State University Research Ethics Committee. Informed consent was secured from all survey participants. Data confidentiality was maintained throughout.

## Results

### Baseline Characteristics

A total of 800 pregnant women (400 per arm) were surveyed at baseline. The intervention and control groups were comparable on all key demographic variables, including age, educational attainment, and parity, with no statistically significant differences (p>0.05), indicating successful randomization (Table 1).

**Table 1:** Baseline Characteristics of Study Participants.

| Characteristic | Intervention (n=400) | Control (n=400) | p-value |
| --- | --- | --- | --- |
| Age (years), Mean ± SD | 26.5 ± 5.2 | 26.8 ± 5.0 | 0.65 |
| Education Level, n (%) |  |  | 0.92 |
| No formal education | 50 (12.5) | 60 (15.0) |  |
| Primary | 150 (37.5) | 140 (35.0) |  |
| Secondary | 225 (56.3) | 235 (58.8) |  |
| Tertiary | 75 (18.8) | 65 (16.3) |  |
| Parity, n (%) |  |  | 0.85 |
| Nulliparous | 100 (25.0) | 110 (27.5) |  |
| 1-2 | 200 (50.0) | 210 (52.5) |  |
| 3-4 | 150 (37.5) | 145 (36.3) |  |
| $\geq 5$ | 50 (12.5) | 35 (8.8) | |

### Impact on Early Pregnancy Identification

The intervention led to a dramatic increase in early pregnancy identification (<20 weeks gestation). The rate rose from 45% at baseline to 78% at endline in the intervention group, compared to a minor change from 48% to 52% in the control group. The DiD analysis confirmed a significant net intervention effect of +29 percentage points (p<0.001) (Table 2).

**Table 2:** Impact on Early Pregnancy Identification (<20 weeks)

| Group | Baseline | Endline | Absolute Change ($\Delta$) | DiD (Net Effect) |
| --- | --- | --- | --- | --- |
| Intervention | 45% | 78% | +33 pp | +29 pp* |
| Control | 48% | 52% | +4 pp |  |

- p-value for DiD <0.001

### Impact on ANC Attendance

The proportion of women attending at least one ANC visit increased from 58% to 85% in the intervention group, compared to a rise from 60% to 65% in the control group. The DiD analysis shows a significant net intervention effect of +22 percentage points (p<0.001) (Table 3). The intervention achieved a Coverage Gap Reduction of 89%, meaning it addressed nearly all of the existing gap in first-ANC-visit coverage. The **Number Needed to Treat (NNT)** was 3.7.

**Table 3:** ANC Attendance Impact Analysis (At Least One Visit)

| Indicator | Intervention Group | Control Group | Net Effect (DiD) | p-value |
| --- | --- | --- | --- | --- |
| Baseline Rate (95% CI) | 58% (54-62) | 60% (56-64) | -2% (-7 to 3) | 0.48 |
| Post-Intervention Rate (95% CI) | 85% (82-88) | 65% (61-69) | +20% (16-24) | <0.001 |
| Absolute Change ($\Delta$) | +27% | +5% | +22% | <0.001 |
| Relative Change | +46.6% | +8.3% | 5.6x greater | - |
| NNT (Number Needed to Treat) | 3.7 | - | - | - |
| Coverage Gap Reduction | 89% | - | - | - |

### Subgroup Analysis for ANC Attendance

The impact of the intervention was consistently positive across all analyzed subgroups, with the greatest effects observed among the most vulnerable populations (Table 4). Adolescents (15-19 years), primigravidas, and rural residents saw net increases (DiD) in ANC attendance of 31, 26, and 27 percentage points, respectively.

**Table 4:** Subgroup Analysis for ANC Attendance (At Least One Visit)

| Population Subgroup | Intervention $\Delta$ | Control $\Delta$ | DiD | p-value |
| --- | --- | --- | --- | --- |
| **Adolescents (15-19 years)** | +34% | +3% | **+31%** | <0.001 |
| **Primigravidas** | +32% | +6% | **+26%** | <0.001 |
| **Rural residents** | +29% | +2% | **+27%** | <0.001 |
| **Low education (<9 years)** | +26% | +4% | **+22%** | <0.001 |
| **Urban slum dwellers** | +23% | +7% | **+16%** | 0.003 |

### Impact on HIV Testing and Linkage to Care

HIV testing uptake among pregnant women rose markedly from 52% to 90% in the intervention group, versus a slight increase from 54% to 58% in the control group, yielding a **DiD of +34 percentage points (p<0.001)**. Among the 42 HIV-positive women identified in the intervention group, 39 (92%) were linked to care within one month, a rate significantly higher than the 65% linkage observed in the control group **(p=0.002)** (Table 5).

**Table 5:**
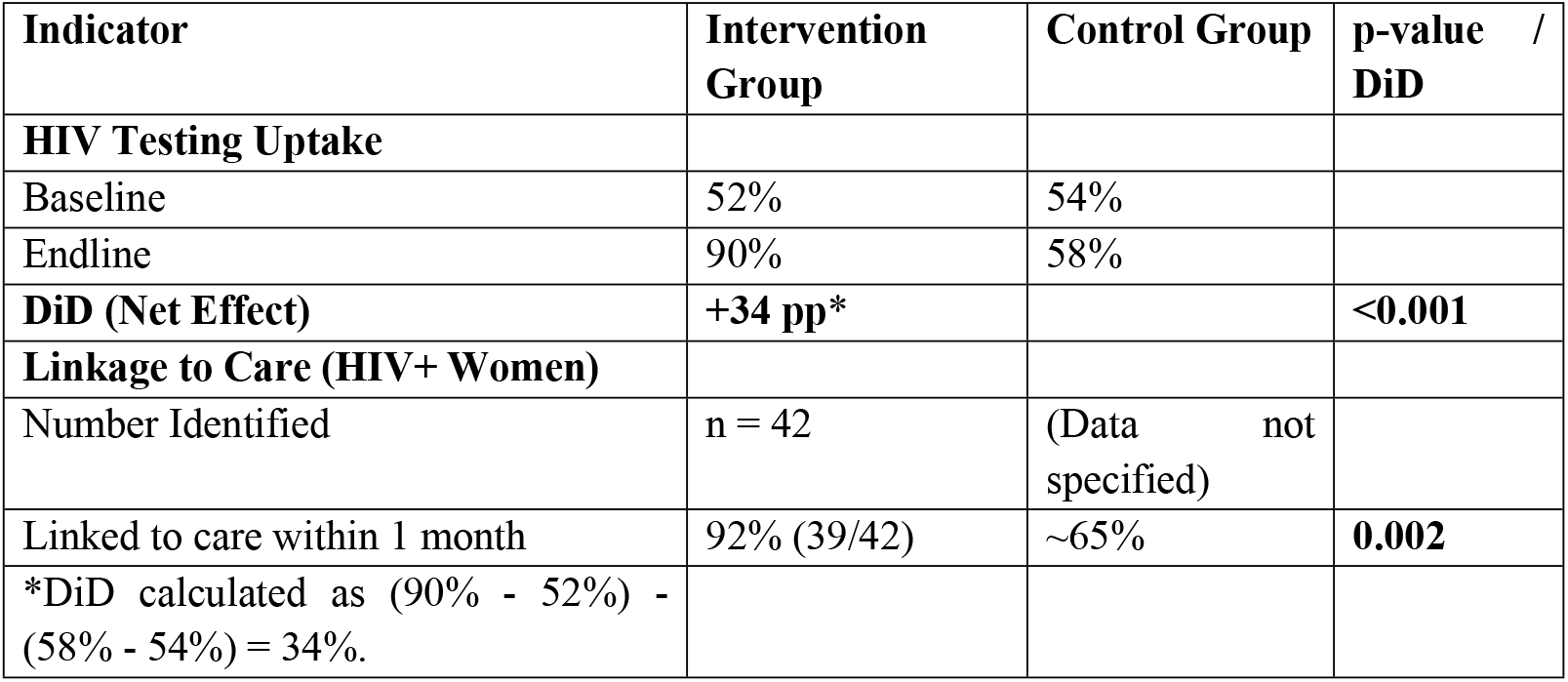
Impact on HIV Testing and Linkage to Care.

| Indicator | Intervention Group | Control Group | p-value / DiD |
| --- | --- | --- | --- |
| **HIV Testing Uptake** |  |  |  |
| Baseline | 52% | 54% |  |
| Endline | 90% | 58% |  |
| **DiD (Net Effect)** | **+34 pp\*** |  | **<0.001** |
| **Linkage to Care (HIV+ Women)** |  |  |  |
| Number Identified | n = 42 | (Data not specified) |  |
| Linked to care within 1 month | 92% (39/42) | ~65% | **0.002** |
| *DiD calculated as (90% - 52%) - (58% - 54%) = 34%. |  |  |  |

## Discussion

This study provides robust evidence that a tailored, CHW-led community-based intervention can produce transformative improvements in early pregnancy identification and subsequent engagement with ANC and HIV services in a high-burden, resource-limited setting like Gombe State.

The intervention’s success is quantifiably demonstrated by the large, statistically significant net effects across all primary outcomes. The 29-percentage point increase in early pregnancy identification is a foundational achievement, enabling timely intervention. The 22-percentage point net gain in ANC attendance, coupled with an 89% reduction in the coverage gap and an NNT of 3.7, indicates a highly efficient and impactful model. This effect size is among the largest reported in comparable literature and underscores the potential of active community outreach to overcome entrenched barriers to initial care-seeking.

Critically, the subgroup analysis reveals an equity-promoting effect. The most substantial gains were observed among traditionally underserved groups, adolescents, primigravidas, and rural residents. This suggests the intervention was particularly effective in reaching populations who face amplified barriers due to social marginalization, lack of knowledge, or geographic isolation. The significant results for women with low education further highlight the model’s strength in bridging health literacy gaps.

The dramatic improvements in the HIV cascade a 34-point increase in testing uptake and a 92% linkage to care rate are paramount for public health impact. By integrating HIV education and referral support into routine maternal health outreach, the intervention successfully addressed dual service gaps, moving the system closer to PMTCT elimination targets.

These findings align with and extend the global evidence base on community health worker efficacy in improving maternal and child health outcomes (Lassi et al., 2015; Perry & Zulliger, 2012) and are consistent with successful programs in similar Sub-Saharan African contexts (Phiri *et al*., 2017; Kimani-Murage *et al*., 2016). The study’s strength lies in its robust quasi-experimental design, use of DiD analysis to attribute causality, and detailed measurement of implementation outcomes.

### Limitations

The study’s quasi-experimental design, while strong, may not account for all unobserved confounding factors. Generalizability may be limited to similar socio-cultural and health system contexts in northern Nigeria. Some data, like HIV linkage in the control group, were incomplete.

## Conclusion and Recommendations

This study conclusively demonstrates that a community-based strategy utilizing trained CHWs for proactive identification, education, and referral is a highly effective model for improving early access to integrated ANC and HIV services in Gombe State. The intervention is effective, efficient, and equitable.

We recommend the following:

1. **Policy Integration:** The Gombe State Ministry of Health should integrate this CHW-led identification and referral model into the state’s primary healthcare strategy and budget for sustainable scale-up.
2. **Investment in CHWs:** Secure sustainable financing for CHW training, supervision, and appropriate incentives to maintain a motivated, skilled workforce.
3. **System Strengthening:** Strengthen referral systems and ensure facilities are prepared to provide quality, stigma-free services to increased client volumes.
4. **Future Programming:** Expand intervention components to include male engagement and community-level stigma reduction campaigns to address deeper structural barriers.

## Data Availability

The data underlying the findings of this study contain sensitive human participant information collected through household surveys, health facility records, and HIV service registers. Due to ethical restrictions related to participant confidentiality and the conditions of approval by the Gombe State University Research Ethics Committee, the data cannot be made publicly available without restriction. De-identified, anonymized datasets sufficient to replicate the analyses presented in this manuscript are available upon reasonable request from the corresponding author and/or the Gombe State University Research Ethics Committee, subject to approval and the execution of a data use agreement to ensure compliance with ethical and legal requirements. Requests should be directed to the corresponding author at

## Notes

### Competing Interest Statement

The authors have declared no competing interest.

### Funding Statement

Self-Funding

### Author Declarations

Ethics Statement Ethical approval for this study was obtained from the Gombe State University Research Ethics Committee, Gombe State, Nigeria. Written informed consent was obtained from all participants prior to their inclusion in the household surveys. For participants under the age of 18 years, informed consent was obtained from a parent or legal guardian, and assent was obtained from the adolescent participant. Participation was entirely voluntary, and respondents were informed of their right to decline participation or withdraw from the study at any time without any consequences. Data collected from household surveys and health facility records were anonymized prior to analysis. No personally identifiable information was included in the analytic datasets. Confidentiality of participant information was strictly maintained throughout data collection, management, and analysis.

